# Built-in Healthcare Applications Reveal Step Changes Associated with Temperature, Transportation, and Marital Status Among Urban Cities in Japan

**DOI:** 10.1101/2025.03.20.25324356

**Authors:** Nobuhiko Wakai, Taiga Yamada, Hiroyuki Tomoyama, Shigehiro Iida

## Abstract

**BACKGROUND:** Walking is a fundamental daily activity representing health status and physical condition. The number of steps taken in a given time period is widely used in research areas such as aging, geriatrics, gerontology, public health, and preventive medicine. However, the underlying mechanisms of step counts are not well understood.

**OBJECTIVES:** To investigate daily step counts associated with temperature, transportation, and marital status.

**DESIGN:** Time series analysis of daily steps using built-in healthcare applications on smartphones.

**SETTING:** Government-designated, well-developed urban cities in Japan: Fukuoka, Kawasaki, Kobe, Kyoto, and Saitama.

**PARTICIPANTS:** Respondents totaled 622 40- to 79-year-olds, comprising 370 males and 252 females.

**MEASUREMENTS:** The mean period of our retrospective data was 2,344 days.

**RESULTS:** Seasonal-trend decomposition using loess was applied to time series steps. With the high coefficient of determination *R*^2^: 0.798, an absolute value function was fitted between temperature and the mean daily steps of the seasonal component. Furthermore, ordinary train usage in Saitama, Kawasaki, and Fukuoka was significantly greater than that in Kobe and Kyoto by 14.1 points (*p* = 0.001). Moreover, married and divorced or bereaved males’ mean daily step counts were significantly larger than those of females’ by 1,832 (*p* = 0.001) and 2,480 (*p* = 0.001), respectively. By contrast, the difference in the mean daily step counts for unmarried males and females was only 100.

**CONCLUSIONS:** This study presents significant associations between mean daily steps and the factors of temperature, transportation, and marital status. These associations can alleviate biases in step research by area and season to facilitate better step count comparisons in many research fields.

## 1. Introduction

### 1.1. Background

Walking is a fundamental daily activity that represents health status [1] and physical condition [2]. Step counts are widely used in various fields such as aging [3], geriatrics [4], gerontology [5], public health [6], and preventive medicine [7, 8, 9]. Step analysis has the advantage of generality; that is, most people walk daily, regardless of age, sex, or residential area. Furthermore, steps are typically measured using non-invasive methods such as pedometers. Analyzing daily steps accurately and over time, however, has remained challenging owing to the need to collect steps in daily life for several contiguous years.

Japan has the most aging population globally; that is, life expectancy at birth is 81.5 years in males and 86.9 years in females as of 2019 [10]. Other high-income developed countries face this societal trend as well because life expectancy in such regions is longer than that in lower-income countries [11]. The World Health Organization (WHO) classifies causes of death into three categories: communicable, non-communicable, and injuries [12]. The non-communicable category tends to impact high-income countries, while low-income countries suffer more from communicable categories (i.e., contagious diseases). To reduce non-communicable causes of death requires people to take care of themselves daily.

Preventive medicine plays a critical role in maintaining the public’s health; its aim is to preserve health by preventing diseases from interrupting our daily lives. The relationships between health and daily step counts have been studied extensively. For example, one meta-analysis [13] reported an inverse association between daily steps and all-cause mortality: a 15% decreased risk of all-cause mortality associated with a 1,000-step increment. Therefore, increasing step counts in daily life is fundamental in preventive medicine. Importantly, however, while the relationship between steps and longevity is important for aging and preventive medicine, walking is sensitive to climates and natural environments, as evidenced by seasonal changes observed in step counts. These changes have historically prevented longitudinal analysis of step changes.

### 1.2. Related work

Large-scale step data have been analyzed using questionnaires or smartphones to improve public health and address aging. From 2009 to 2019, the Japanese government annually conducted one of the largest step surveys in Japan with 4,591 participants, which included questions about physical activities and nutrition [14]. This survey reported that the mean daily step counts were 6,793 and 5,832 for males and females, respectively. Unfortunately, the survey was discontinued because of the COVID-19 pandemic. Some step studies, such as the survey mentioned above, use pedometers to record daily steps, while others use smartphone applications for walking [15, 16, 17] and tracking physical activity [18]. Importantly, smartphone applications can allow for greater numbers of participants because researchers are not required to provide pedometers.

Seasonal changes in daily step counts have also been examined in Japan because aging is a longitudinal process analyzed using long-term data; however, seasonal changes affect long-term step counts. Although previous work has accounted for seasonal changes, the participants were all in their 70s, and the sample sizes were only 41 [19], 39 [20], and 22 [21]. These studies were pioneering but inadequate because of the lack of quantitative analysis [19] and discrete time-series steps. The intervals of data collection were 3 months and thus did not reflect the continuous four seasons [20, 21]. As described above, seasonal changes in step counts were investigated throughout the year; however, these studies did not disentangle factors such as age, residential areas, and climate. Such disentangling has remained an open challenge because of the lack of long-term step data and reliable participants.

### 1.3. Our approach

To address the above-mentioned issues, we obtained step data that were automatically stored on smartphones. These data were collected from 622 residents, aged 40 to 79 years, from five urban cities in Japan. This is the first study that measured daily steps using time series methods from before the COVID-19 pandemic to after. Using time series step data, we obtained a regression line between daily steps and outside temperature. Furthermore, we analyzed the mean daily steps associated with transportation modes to explore the differences among cities. Additionally, we compared the impact of marital status on mean daily steps. The purpose of this study was to investigate the associations between daily step counts and temperature, transportation, and marital status.

## 2. Methods

### 2.1. Study cities

Previous step studies have been conducted in a city [16, 17, 19, 20, 21], a county [14], and multiple countries [15]; however, they did not report the impact on daily steps of differences among cities. It is important to understand these differences because public health and city planning often focus on city communities. Therefore, we selected five cities in Japan: Fukuoka City, Fukuoka Prefecture; Kawasaki City, Kanagawa Prefecture; Kobe City, Hyogo Prefecture; Kyoto City, Kyoto Prefecture; and Saitama City, Saitama Prefecture. These cities have humid subtropical climates with four distinct seasons, but it rarely snows in winter. The geographical features of the cities primarily comprise flat land. The five cities were selected for comparability independent of climate and developing conditions. That is, given the similar climates and development, the focus was on differences in transportation.

We selected government-designated cities in Japan—cities that possess equal authority over prefectures—to create alignment in economic development across the sample and thereby minimize potential obstacles that might prevent residents from walking or being active outdoors. The Japanese government enables cities with populations of 500,000 or more, excluding Tokyo, to have swift administrative responses because some prime cities are comparable to prefectures as indicated in the local autonomy act [22]. These cities have well-developed infrastructure: streets, sidewalks, bridges, parks, public facilities, and public transportation. Twenty prime cities in Japan were designated as of December 26, 2024. The populations of the five selected cities in this study, as shown in Table 1(a), were continuously ranked from fifth to ninth in the list of 20 government-designated cities. Thus, these cities have approximately equal populations. Furthermore, mean temperatures for the four seasons differ less than 3*^◦^*C among the five cities. The distance between a study city and the nearest large cities—government-designated cities or the 23 wards of Tokyo—using city locations [23], as shown Table 1(a) because cities are connected with transportation such as trains and cars.

**Table 1: (a).** Demographics of study cities.

|  | Fukuoka | Kawasaki | Kobe | Kyoto | Saitama |
| --- | --- | --- | --- | --- | --- |
| Population ($10^3$) [31] | 1612 | 1538 | 1525 | 1464 | 1324 |
| Area ($\text{km}^2$) [37] | 343.47 | 142.96 | 556.93 | 827.83 | 217.43 |
| Distance between cities (km)* | 54.6 | 3.6 | 28.1 | 42.8 | 13.5 |
| Location (degree) [23] |  |  |  |  |  |
| Latitude | 33.590 | 35.531 | 34.689 | 35.012 | 35.862 |
| Longitude | 130.402 | 139.703 | 135.196 | 135.768 | 139.646 |
| Mean temperature ($^{\circ}\text{C}$) [38] | | | | | |
| Spring (March–May) | 16.4 | 15.5 | 15.7 | 15.3 | 14.8 |
| Summer (June–August) | 27.1 | 25.6 | 26.7 | 26.9 | 25.8 |
| Autumn (September–November) | 20.1 | 19.1 | 20.1 | 19.0 | 18.0 |
| Winter (December–February) | 8.4 | 7.8 | 7.6 | 6.3 | 5.7 |

\* Distance between a study city and the nearest government-designated cities or the 23 wards of Tokyo [23].

**Table 1: (b).**
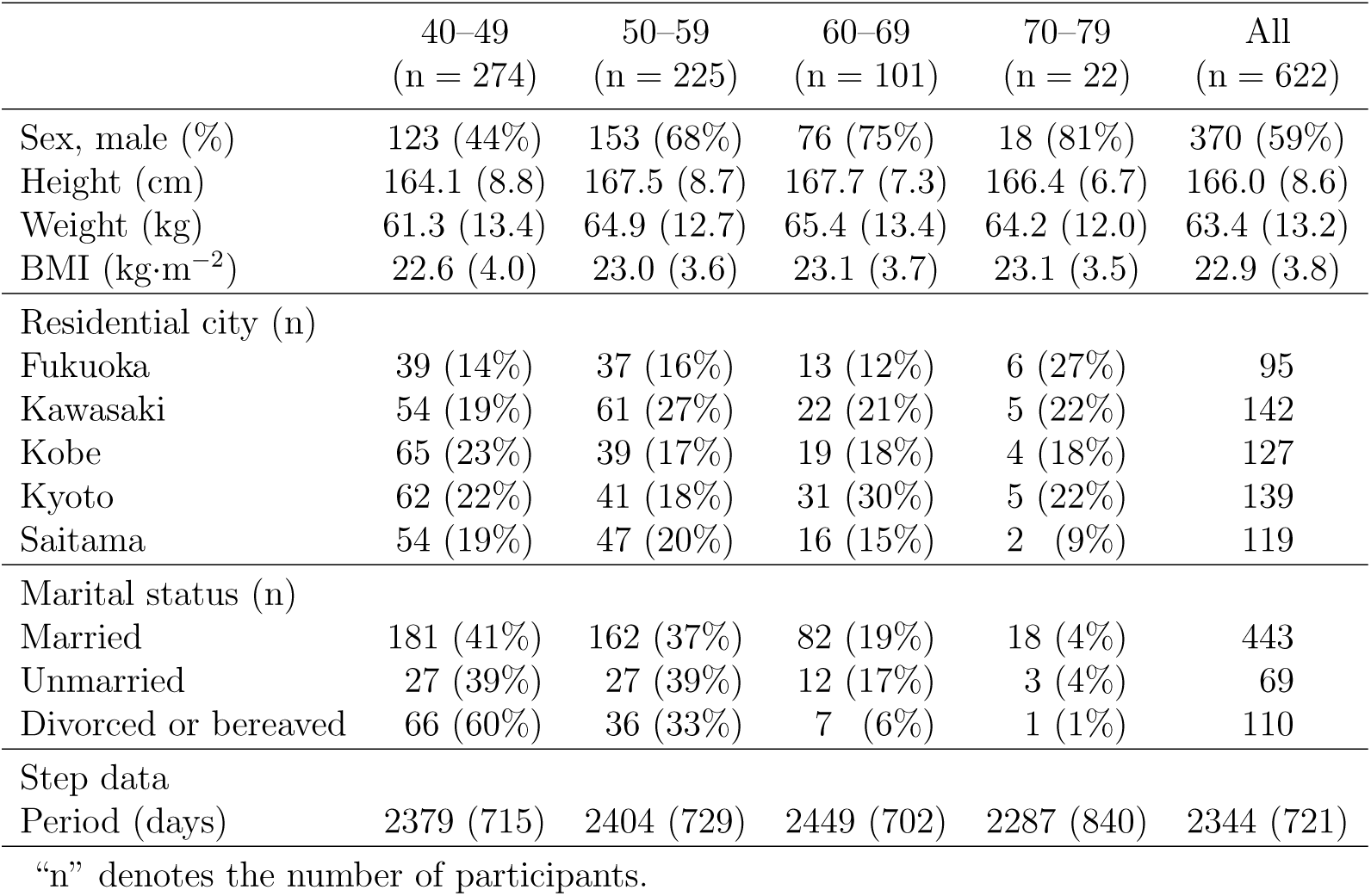
Demographics of study participants stratified by age as mean (standard deviation) or counts (%)

|  | 40–49<br>(n = 274) | 50–59<br>(n = 225) | 60–69<br>(n = 101) | 70–79<br>(n = 22) | All<br>(n = 622) |
| --- | --- | --- | --- | --- | --- |
| Sex, male (%) | 123 (44%) | 153 (68%) | 76 (75%) | 18 (81%) | 370 (59%) |
| Height (cm) | 164.1 (8.8) | 167.5 (8.7) | 167.7 (7.3) | 166.4 (6.7) | 166.0 (8.6) |
| Weight (kg) | 61.3 (13.4) | 64.9 (12.7) | 65.4 (13.4) | 64.2 (12.0) | 63.4 (13.2) |
| BMI ($\text{kg}\cdot\text{m}^{-2}$) | 22.6 (4.0) | 23.0 (3.6) | 23.1 (3.7) | 23.1 (3.5) | 22.9 (3.8) |
| Residential city (n) |  |  |  |  |  |
| Fukuoka | 39 (14%) | 37 (16%) | 13 (12%) | 6 (27%) | 95 |
| Kawasaki | 54 (19%) | 61 (27%) | 22 (21%) | 5 (22%) | 142 |
| Kobe | 65 (23%) | 39 (17%) | 19 (18%) | 4 (18%) | 127 |
| Kyoto | 62 (22%) | 41 (18%) | 31 (30%) | 5 (22%) | 139 |
| Saitama | 54 (19%) | 47 (20%) | 16 (15%) | 2 (9%) | 119 |
| Marital status (n) |  |  |  |  |  |
| Married | 181 (41%) | 162 (37%) | 82 (19%) | 18 (4%) | 443 |
| Unmarried | 27 (39%) | 27 (39%) | 12 (17%) | 3 (4%) | 69 |
| Divorced or bereaved | 66 (60%) | 36 (33%) | 7 (6%) | 1 (1%) | 110 |
| Step data |  |  |  |  |  |
| Period (days) | 2379 (715) | 2404 (729) | 2449 (702) | 2287 (840) | 2344 (721) |

“n” denotes the number of participants.

### 2.2. Data collection and study participants

We obtained step data using a built-in healthcare application in Apple iPhone smartphones to analyze time series step counts for several years. Apple’s step-counting algorithm is proprietary, and the details are not disclosed. However, steps estimated by Apple iPhone smartphones corresponded to pedometers at various speeds: slow, self-selected, and fast, with the coefficient of determination at 0.98, 0.98, and 0.99, respectively [24]. The healthcare application is active by default and automatically stores steps counted by a part of the operating system using smartphone acceleration sensors. The steps are recorded with standard time stamps, and we could aggregate the steps using arbitrary periods, such as steps per day. Furthermore, healthcare data are recorded even if users change phone models, as long as user accounts are not changed.

To minimize selection bias, we recruited extensively (via the internet), between April 28, 2024 and October 21, 2024, people who always carry their Apple iPhone smartphones. Our retrospective data consisted of adults aged 40 to 79 years living in the five selected cities in Japan. Although we only recruited iPhone smartphone users, the potential selection bias was negligible: as of 2021, 95% of Japanese people aged 13 to 69 years use smartphones, and Apple iPhone dominated 46.6% market share based on the number of devices in Japan [25]. We believe this popularity produces limited bias of iPhone users compared with Android smartphone users or people who do not use smartphones. We also obtained the following participant information to complete our analysis: age, sex, height, weight, residential areas, marital status, and daily transportation usage.

### 2.3. Demographic characteristics

To the best of our knowledge, we are the first researchers to directly analyze step data stored in the built-in healthcare application on Apple iPhone smartphones. We employed data cleansing empirically to eliminate outliers because of the lack of de facto cleansing procedures. The participants’ demographics, after data cleansing, are shown in Table 1(b). The 622 participants comprised 370 males and 252 females aged 40 to 79 years. The number of participants in each city was at least 95. The mean period of data collection among all participants was 2,344, encompassing repeated seasons.

For the data cleansing process, we first aggregated daily steps using each participant’s healthcare data. We sliced the time-series daily steps with the period between January 1, 2016, and May 26, 2024, because participants’ recordings covered various periods. The beginning date was selected to capture an adequate number of participants: 196 people began their step recordings on January 1, 2016. The end date of the slicing corresponded to the earliest data submission date among recruited participants. Second, we removed any data showing zero daily steps; this likely indicated that the participant had forgotten to carry their phone with them (we had previously confirmed that participants always carried their smartphones throughout the questionnaires). Third, we applied the Smirnov-Grubbs test [26] to each participant. This test excluded outlier steps using a 5% significance level. Finally, we calculated the mean *µ* and standard deviation *σ* for the mean daily steps of all participants, and then eliminated 36 participants whose mean daily steps exceeded the threshold of *µ* + 1.645*σ*: 13,750 daily steps. This threshold corresponded to the upper 5% participants on the normal distribution assumption.

### 2.4. Statistical analysis

We considered the period of the COVID-19 pandemic as part of the research period because the pandemic’s impact was notable enough to affect analysis of the time series steps over a long period. COVID-19 cases in Japan trended in seven waves, each displaying a rapid increase in cases [27]. The Japanese government officially implemented measures in response to the COVID-19 pandemic, and although they did not enforce lockdowns, citizens were strongly recommended to obey the declaration of the state of emergency on April 7, 2020, and the quasi-state of emergency on April 21, 2022. These declarations suppressed activities and transportation (e.g., outings and public events), thus shortening restaurant business hours and reducing school and company commutes. Following [28], and on the basis of the two declarations, we defined the period of the COVID-19 pandemic in Japan as April 7, 2020 to April 21, 2022. Hence, the numbers of participants before and after the COVID-19 pandemic were 541 and 622, respectively.

Unlike a previous step analysis [18], our collected step data included factors such as time series, city, age, and sex. To examine these steps, we first focused on the time series step counts to process long-term trends for other comparisons. It was essential to extract long-term trends because of the COVID-19 pandemic. This extraction was conducted with Seasonal-Trend decomposition using Loess (STL) [29]. The STL is one of the filtering procedures for decomposing a time series into trend, seasonal, and remainder components. For a yearly periodicity, we used 365 days for the number of observations *n*_(_*_p_*_)_ in each cycle of the seasonal component. Standard STL parameters were used for the other factors in daily steps: *n*_(_*_i_*_)_ = 5, *n*_(_*_o_*_)_ = 0, *n*_(_*_l_*_)_ = 367, *n*_(_*_s_*_)_ = 7, and *n*_(_*_t_*_)_ = 1.5 *· n*_(_*_p_*_)_*/*(1 *−* 1.5*/n*_(_*_s_*_)_). The input for STL was a time series of the mean daily steps among participants.

Second, we analyzed the seasonal component of the STL result associated with temperature. A regression line was calculated between the mean daily steps and *| Temperature − T_opt_ |*, where *T_opt_* represents the optimal temperature, to maximize the coefficient of determination *R*^2^. Third, we compared mean daily steps across the five study cities after the COVID-19 pandemic. Furthermore, we examined ordinary transportation usage to compare two groups: cities with higher mean daily steps and those with lower mean daily steps. Finally, we focused on the marital status of the participants. Using the questionnaire, we classified the participants into three categories: married, unmarried, and divorced or bereaved. In the *t*-test and *z*-test, we used the Hedges’ effect size *g* to evaluate differences [30].

## 3. Results

### 3.1. Time series step analysis

We analyzed time series daily steps throughout the period between January 1, 2016, and May 26, 2024, to observe year-order trends. The results from STL using all participants are shown in Figure 1. The trend component observed an increased tendency from 5,551 to 7,105 steps from August 16, 2020, to May 26, 2024. The remainder had large deviations caused by large-scale events, such as typhoons and the declarations of the state or quasi-state of emergency. Because the trend and remainder components extracted non-periodic step deviations, the seasonal component had zero-center periodic deviation within about *±*1000 steps, representing seasonal changes. Although the seasonal component had noise, the 30-day moving average indicated apparent periodicity, as shown in Figure 1.

**Figure 1:**
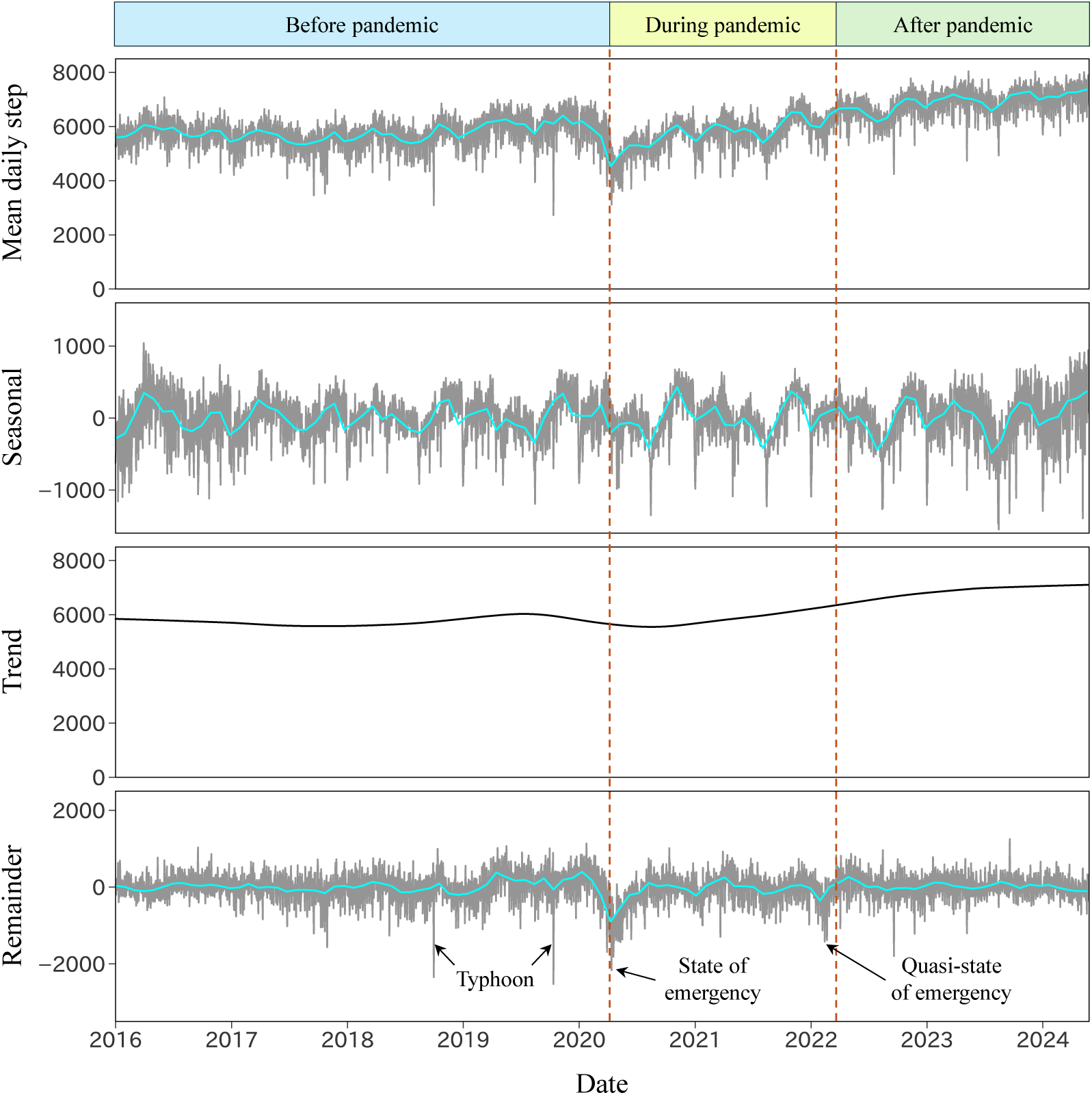
Results of STL for mean daily steps among participants throughout all time periods. From top to bottom: input daily steps and the seasonal, trend, and remainder components. Cyan lines represent 30-day moving averages. Two dashed brown lines indicate the beginning and end dates of the COVID-19 pandemic on April 7, 2020, and March 21, 2022, respectively. The remainder component has peaks caused by typhoons and the declarations of the state or quasi-state of emergency.

To determine seasonal changes, we further examined the STL results. After calculating the mean daily steps of seasonal components in months, we obtained M-shaped changes associated with the mean daily steps and months in Figure 2(a). The highest mean daily step count for the seasonal component was 282 steps in November, the mean temperature of which was 13.7*^◦^*C. By contrast, the lowest mean daily step count was *−*328 steps in August, with a mean temperature of 28.7*^◦^*C, which corresponded to the highest temperature in a year. Step changes in the four seasons were also M-shaped; however, the highest and lowest mean daily step counts did not correspond to the mean daily step counts in months. A regression line between the mean daily steps of seasonal components in months, *y*, and *| Temperature − T_opt_ |*, *x*, was *y* = *−*34.44*x* + 246.59 with the coefficient of determination *R*^2^: 0.798, as shown in Figure 2(b), and the 14.3*^◦^*C of the optimal temperature *T_opt_* maximized the *R*^2^.

**Figure 2:**
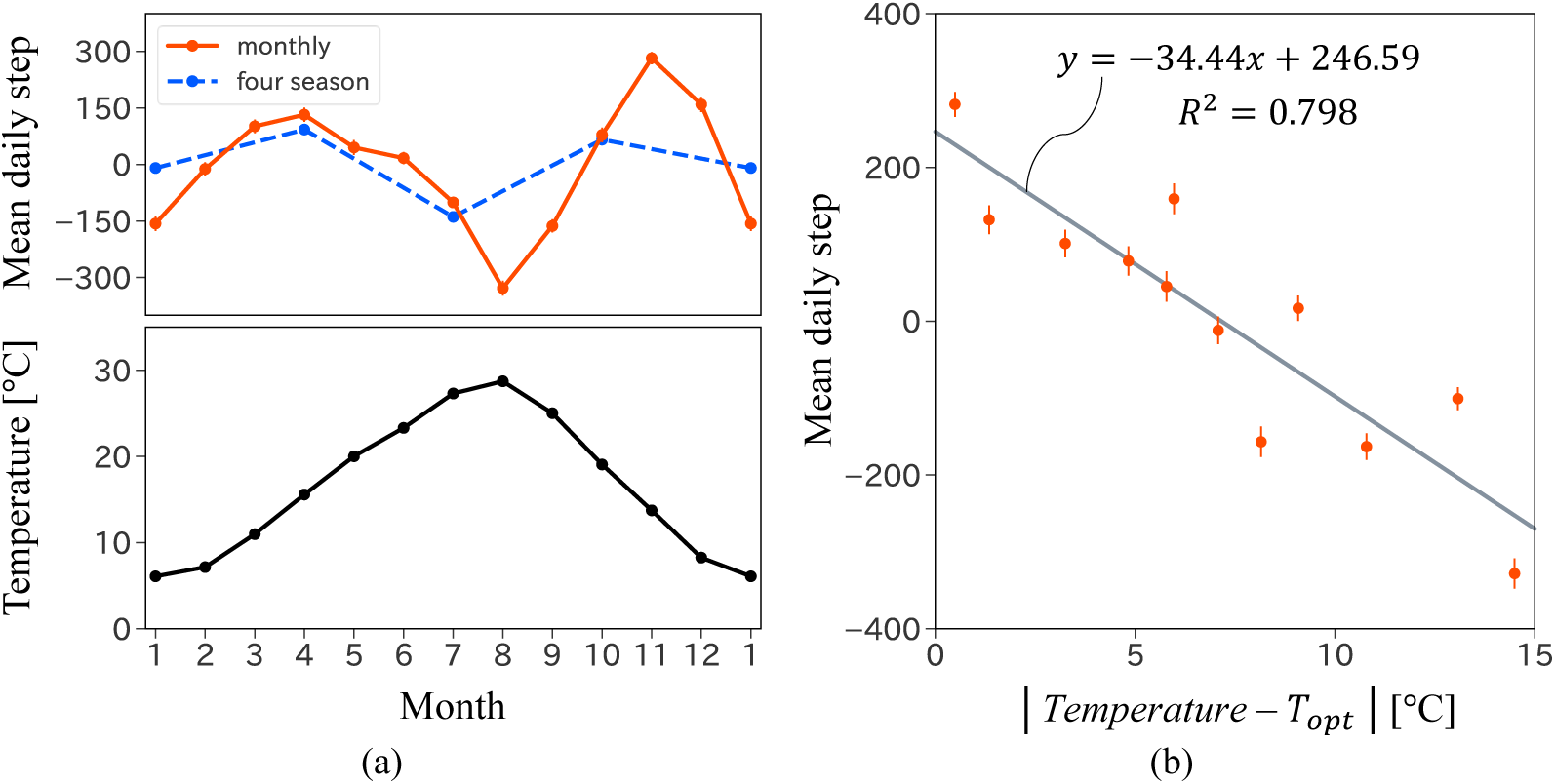
Seasonal changes in mean daily steps using the STL seasonal component results. The error bars are standard errors. (a) The top is the mean daily steps in months. Red and blue lines indicate the mean daily steps for the months and four seasons, respectively. The four seasons are defined as follows: spring (March–May), summer (June–August), autumn (September–November), and winter (December–February). The bottom represents the mean temperature in months. (b) Mean daily steps for the seasonal component in months depend on *| Temperature − T_opt_ |*, where *T_opt_* is 14.3*^◦^*C. The gray line indicates the regression line.

### 3.2. Step comparison among cities

To investigate differences among the cities, we compared the mean daily step counts after the COVID-19 pandemic because mean daily step counts slumped during the pandemic, as shown in Figure 1. Mean daily step counts were 7493*±*331, 7439*±*394, 7238*±*354, 6407*±*277, and 5920*±*300 (mean*±*SE) in Saitama City, Fukuoka City, Kawasaki City, Kobe City, and Kyoto City, respectively, as shown in Figure 3(a). Significant differences were observed, using a two-sided *t*-test, in pairs of cities (*p*-value, Hedges’ *g*): Saitama and Kyoto (*p* = 0.001, *g* = 0.441), Fukuoka and Kyoto (*p* = 0.001, *g* = 0.415), Kawasaki and Kyoto (*p* = 0.003, *g* = 0.339), Saitama and Kobe (*p* = 0.006, *g* = 0.322), and Fukuoka and Kobe (*p* = 0.014, *g* = 0.299). On the basis of the significant differences in mean daily step counts, the cities were classified into two categories: higher and lower. The higher cities were Saitama, Fukuoka, and Kawasaki, and the lower cities were Kobe and Kyoto. Their mean daily steps were 7377*±*208 and 6153*±*205 (mean*±*SE) for higher and lower cities, respectively, and the 1,224-step difference in mean daily steps was significant (*p* = 0.014, *g* = 0.178).

**Figure 3:**
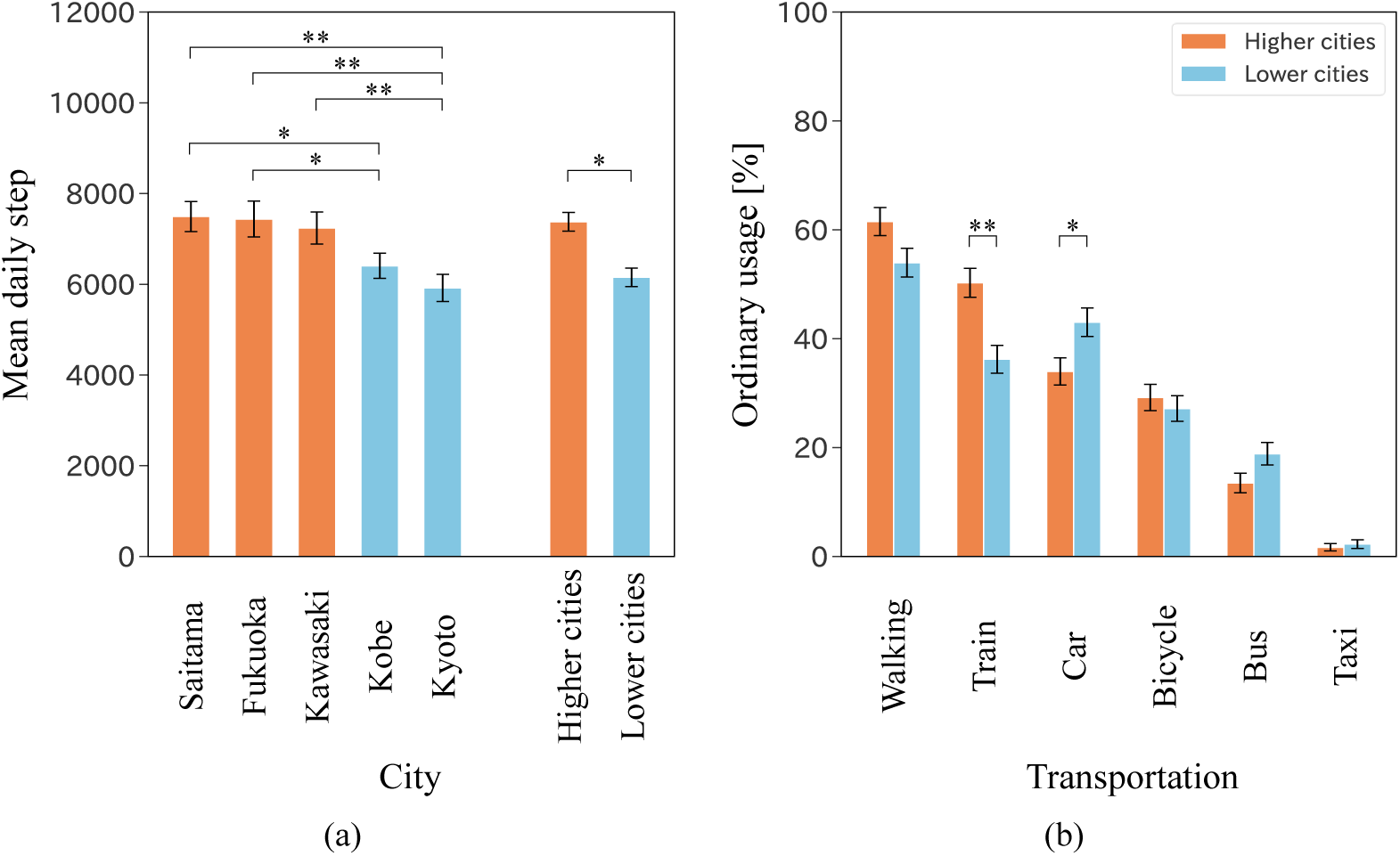
City comparison of mean daily steps after the COVID-19 pandemic. The error bars are standard errors. The * and ** indicate the significant difference of 5% and 1% significance levels, respectively. (a) The mean daily step counts for the study cities are indicated from the first to fifth bars. The last two bars on the right represent the aggregated cities: the higher cities consist of Saitama, Fukuoka, and Kawasaki, and the lower cities consist of Kobe and Kyoto. (b) Ordinary transportation usage is shown. This usage is based on the participant’s questionnaires, and multiple answers are permitted. The higher cities and lower cities correspond to the definition of (a).

We also examined the association between daily steps and transportation modes to clarify the difference between the higher and lower cities. Figure 3(b) shows ordinary transportation usage: walking, trains, cars, bicycles, buses, and taxis. The questionnaire allowed participants to select multiple answers for transportation mode. The results showed two significant differences using a two-sided *z*-test. The ordinary usage of trains in the higher cities was significantly larger than that in lower cities by 14.1 points (*p* = 0.001, *g* = 0.283). Additionally, the ordinary usage of cars in the higher cities was significantly smaller than that in lower cities by 9.0 points (*p* = 0.011, *g* = 0.186).

The train usage and the distance from the nearest large cities in the higher cities were as follows (usage, distance): Saitama City (55.4%, 13.5 km), Kawasaki City (57.0%, 3.6 km), and Fukuoka City (33.7%, 54.6 km). Although Fukuoka City is more isolated, Saitama City and Kawasaki City had comparatively high train usage and short distances. By contrast, train usage and the distances for lower cities were as follows: Kobe City (47.2%, 28.1 km) and Kyoto City (26.1%, 42.8 km). Overall, the higher cities had more train usage and shorter distances from the nearest large cities.

### 3.3. Step comparison for marital status

We also studied daily step counts as a function of marital status after the COVID-19 pandemic because daily steps tended to slump during the pandemic, as illustrated in Figure 1. The mean daily step counts associated with marital status are shown in Figure 4. Using a two-sided *t*-test, we observed that the mean daily step counts of married and divorced or bereaved males were significantly greater than those for females of similar status by 1832 (*p* = 0.001, *g* = 0.490) and 2480 (*p* = 0.001, *g* = 0.761), respectively. By contrast, the mean daily steps of unmarried males and females were 6705*±*528 and 6605*±*383 (mean*±*SE), respectively, and the 100-step difference was non-significant. Additionally, the mean daily steps of all participating males were significantly greater than those of females by 1649 (*p* = 0.001, *g* = 0.453), which was similar to the results for married participants because 443 (71%) participants were married, as shown in Table 1(b).

**Figure 4:**
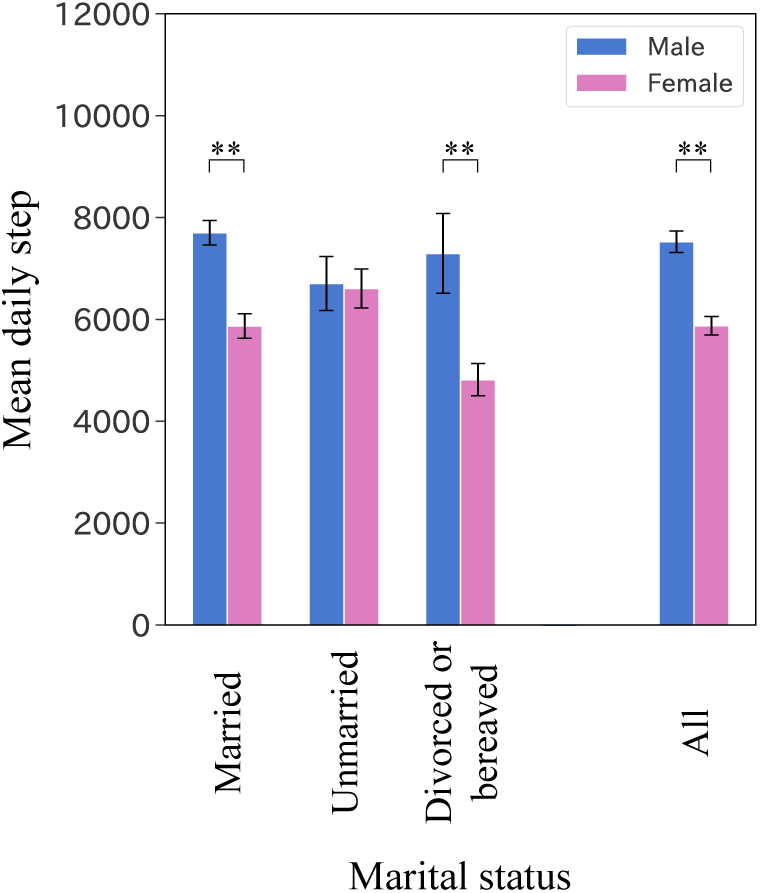
Marital status comparison of mean daily steps after the COVID-19 pandemic. The error bars are standard errors. The ** indicates the significant difference of 1% significance level.

## 4. Discussion

### 4.1. Verifying established steps

To analyze differences in step data across measuring methods, we compared mean daily steps in Japan. In a 3,815-participant survey conducted by the Japanese government in 2019 [14], pedometers were used to collect data from participants who were 40 to 79 years old; daily steps for males, females, and all participants were, respectively, 6518*±*4248, 5649*±*3655, and 6054*±*3942 (mean*±*SD). In the present study, we used built-in healthcare applications on smartphones to measure step counts of participants aged 40 to 79 years after the COVID-19 pandemic; daily steps for males, females, and all participants were, respectively, 7524*±*4067, 5874*±*2910, and 6855*±*3729. The mean daily steps of all participants for the 2019 survey were smaller than those of our measurement by 801 daily steps. This difference suggests that well-developed infrastructure in government-designated cities stimulates more walking and outings, compared with the infrastructure in the cities surveyed throughout Japan. In addition to pedometers, we compared daily steps measured with the Azumio Argus smartphone application [18]. Measurements using the smartphone application, collected between July 2013 and December 2014 in Japan, indicated that the mean daily steps were 6,010 among the participants who used the application for at least 10 days. Similar to the government survey [14], the daily steps measured with the smartphone application were collected throughout Japan. As a result, the mean daily steps for this measurement were smaller than our measurement by 845 daily steps.

Furthermore, we assessed body mass index (BMI) according to sex compared with the Japanese government survey [14]. That survey reported BMIs of 24.1*±*3.3, 22.7*±*3.4, and 23.3*±*3.4 (mean*±*SD), respectively, for males, females, and all participants (aged 40 to 79 years). We observed similar BMIs in the present study: 23.8*±*3.4, 21.5*±*4.0, and 22.9*±*3.8 for males, females, and all participants, respectively.

As discussed above, our measurements obtained using the built-in healthcare application had no contradictions in the mean daily steps aggregated across all participants. In step collection, we cannot escape the trade-off between the length of study periods and the number of participants. Questionnaire surveys and non-built-in smartphone applications can reach many participants; however, these approaches capture only limited time ranges, which comprise the study periods. That is, retroactive step counts cannot be collected before the commencement of a study. By contrast, using built-in healthcare applications has the advantage of obtaining long-term and accurate step counts comparable to pedometers. Although using the built-in healthcare application is time-consuming, only our retrospective data had continuous time-series steps over several years. Therefore, our approach is superior to conventional approaches that use pedometers and non-built-in smartphone applications, which were typically used in previous work.

### 4.2. Comparing seasonal changes

We obtained a regression line between the mean daily steps of STL seasonal components, as shown in Figure 2(b). Because STL extracts trends and noise, the coefficient of determination *R*^2^ was 0.798, indicating a well-fitted association compared with previous work using older adults.

In gerontology, the step counts of older adults in Japan were measured as a function of seasonal changes [19, 20, 21]; seasonal changes in step counts were reported in combination with temperature and length of day [19]. This continuous measurement throughout a year was conducted using pedometers attached to 41 older adults (aged 71*±*4 years) in Nakanojo Town, Gunma, Japan. Although seasonal changes were observed—as presented only in various figures as plots—the quantitative difference between seasons was not reported. To analyze differences among seasons, step counts were collected from 39 older adults (aged 70.7*±*3.2 years) in Kahoku City, Ishikawa, Japan, from Summer 2005 to Winter 2008 [20]. In the summer-to-winter transition, daily steps were 8084*±*3237 in summer and 6098*±*2625 in winter. In another study, the daily steps of 22 older adults (aged 75.1*±*7.3 years) were analyzed in Gero City, Gifu, Japan [21]. The daily steps recorded in spring (6242*±*3229) were significantly higher than those recorded in winter (4918*±*3173).

Each of these three studies [19, 20, 21] reported seasonal changes as fewer daily steps in winter; however, seasons and steps were inconsistent among the studies. For example, the data collection procedures indicate that location and time periods differed in important ways. Gero City is a mountain district, but Nakanojo Town and Kahoku City are flatlands. Furthermore, these three areas have smaller populations than government-designated cities. Specifically, the populations in Nakanojo Town, Kahoku City, and Gero City are 15,386, 34,889, and 30,428, respectively [31]. Therefore, the geographical features and extent of development vary among these areas. Consequently, we cannot interpret seasonal changes in steps as a simple association that aligns with previous studies [19, 20, 21].

Although a quadratic function between daily steps and temperature was reported [19], the number of participants and the observation period were inadequate to reveal the details of the association, yielding a small coefficient of determination *R*^2^: 0.318. By contrast, we obtained the association between daily steps and mean temperature as an absolute value function with a high *R*^2^: 0.798. Our clear association was derived from STL for time series steps. This STL alleviated year-order trends and the deviation of daily steps caused by specific events, such as typhoons and the declaration of a state of emergency. Our association between steps and mean temperature indicated that 14.3*^◦^*C is the optimal temperature *T_opt_*: comfortable conditions for walking and outings. The mean temperature of 14.3*^◦^*C approximately corresponds to spring and autumn in our study cities, and its daytime temperature is approximately 20*^◦^*C to be suitable for walking and outings. Although participants in our study comprised various ages and sexes, the absolute value function *| Temperature − T_opt_ |* was commonly the optimal temperature for them. We interpreted the single optimal temperature as similar to the optimum room temperature for a variety of people; that is, the preferable temperature is generally independent of age and sex. Therefore, our regression line enables us to alleviate the step-related biases of temperature for comparisons across various areas and seasons. This bias reduction will be helpful for vast research fields such as aging, geriatrics, gerontology, public health, and preventive medicine to facilitate better step count comparisons.

### 4.3. Association between steps and transportation

Mean daily steps differed significantly between pairs of cities in the present study, despite the selection of government-designated cities with similar populations. As noted earlier, the cities were classified into higher and lower cities according to their mean daily steps, as shown in Figure 3(a). Participants from the higher cities tended to use trains and not use cars, as shown in Figure 3(b). These results indicate that ordinary transportation usage affects mean daily step counts.

One important factor in increasing daily steps is the city environment [18]. To quantitatively assess city environments, city environments in the entire US and Canada were assessed using the Walk Score [32], which represents the extent of the built environment in relation to walking routes to destinations such as grocery stores, schools, parks, restaurants, and retail centers. City environments in Japan have also been assessed, using the Japanese walkability index [33]. However, the Japanese walkability index in our study cities was not provided in [33], and the index does not include transportation.

We focused on transportation for our city comparisons because we assume that the government-designated cities are equally developed and provide adequate amenities. Public transportation in Japan is popular among people of various ages, sexes, and income levels because of its accurate schedules, safety, and reasonable prices. Train usage data and the distances from the nearest large cities suggest that neighboring large cities induce residents to visit them by train. Unlike cars, buses, and taxis, trains do not deliver people to their final destinations, which leads to increased daily steps. Saitama City and Kawasaki City (two of the higher cities) are located near Tokyo, the capital of Japan. Train usage for Saitama City and Kawasaki City in the present study was 55.4% and 57.0%, respectively, and many residents in the two cities probably go shopping and commute to Tokyo by train. By contrast, train usage in Kyoto City was only 26.1%, and the mean daily step count was the smallest among the study cities because the distance between Kyoto City and its nearest large city is 42.8 km, which is too long a distance for daily transportation. Furthermore, Kyoto City is one of the oldest cities in Japan; it was established as an ancient capital in 794 before trains and cars were invented, and it has maintained its community and urban structure, even avoiding severe damage throughout World War II. This history implies that residents in Kyoto City tend to go shopping near their houses because they can live comfortably within a small area. Therefore, our findings highlight the role of trains in promoting mean daily steps in Japan, and provide potential insights for preventive medicine and urban planners.

### 4.4. Association between steps and marital status

In our comparison of daily steps and their dependence on marital status, we found that married males reported significantly more daily steps than married females, as shown in Figure 4. By contrast, the difference in mean daily steps between unmarried males and females was nonsignificant.

Similar to our results, previous studies [14, 18] have reported that males’ mean daily steps exceeded those of females. Walking is a low-intensity exercise and one of the most fundamental daily activities. On the basis of these features then, we assume that the step-count differences seen in marital status were primarily caused by lifestyle variations. In general, unmarried people spend more of their time working and performing their daily lives, and the lifestyle difference between males and females in Japan is small [34]. By contrast, the lifestyle of married females in Japan changes; for example, 28.8% of them resigned from work, and 6.9% of them switched from regular employment to non-regular employment after marriage [35] for parenting and dedicating themselves to their family [34]. Additionally, Figure 4 suggests that divorced or bereaved people do not return to their unmarried lifestyles in terms of mean daily steps. Therefore, although the results are preliminary, our exploration underscores the importance of considering marital status when analyzing or comparing daily step counts.

### 4.5. Limitations

While this study offers valuable insights, inherent limitations were imposed by data collection using built-in healthcare applications, by the limited number of participants and cities, and by the limited questionnaire. Some of the step data collected from the built-in healthcare application was missing, despite having selected participants who reportedly always carry smartphones. However, we found no contradictions in our comparisons with previous work regarding our step data and that of other studies. The number of participants and cities was limited because data collection was time-consuming. Future studies should validate our results using different cities and/or countries. While our questionnaire explored several important factors relating to step differences, many other factors that may affect daily steps were not considered. We believe that subsequent studies can extend this work to disentangle the details of the mechanisms underlying step count differences and changes.

## 5. Conclusion

This study presents significant associations between mean daily step counts and the factors of temperature, transportation, and marital status among a large sample of urban citizens in Japan. Time series step data were collected, before and after the COVID-19 pandemic, using a built-in healthcare application on smartphones. Our findings highlight the trend of step increase and seasonal changes. These changes associated with temperature can alleviate biases in step research by area and season to facilitate better step count comparisons in many research fields such as aging, geriatrics, gerontology, public health, and preventive medicine. In our comparison of cities, we found that transportation environments significantly affect residents’ daily step counts. This finding underscores the importance of the built environment, which includes not only sidewalks and parks but also transportation modes, to promote daily steps for health. Moreover, our findings on the association between daily steps and marital status can help us understand some differences between males and females that are relevant for subsequent step studies.

## Additional information

### Ethics declarations

The study conformed to the Ethical Guidelines for Medical and Biological Research Involving Human Subjects [36] based on the 1964 Declaration of Helsinki with its subsequent amendments, notified by the Japanese government. Data handling and analysis were approved by the Digital Transformation & Cyber-Physical Systems Division, Panasonic Holdings Corporation. Agreement to participate was obtained from all participants prior to study commencement.

### Availability of data and materials

Not applicable.

### Author contributions

Study conceptualization and design: NW, TY, and SI; methodology and formal analysis: NW and TY; survey: NW, TY, and HT; writing–original draft preparation: NW; writing–review and editing: TY, HT, and SI; Supervision: SI.

## Data Availability

Not applicable.

